# Myocyte Specific Upregulation of *ACE2* in Cardiovascular Disease: Implications for SARS-CoV-2 mediated myocarditis

**DOI:** 10.1101/2020.04.09.20059204

**Authors:** Nathan R. Tucker, Mark Chaffin, Kenneth C. Bedi, Irinna Papangeli, Amer-Denis Akkad, Alessandro Arduini, Sikander Hayat, Gökcen Eraslan, Christoph Muus, Roby Bhattacharyya, Christian M. Stegmann, Human Cell Atlas Lung Biological Network, Kenneth B. Margulies, Patrick T. Ellinor

**Author notes:** **Corresponding Author:** Patrick T. Ellinor, MD, PhD, The Broad Institute of MIT and Harvard, 75 Ames Street, Cambridge, MA 02142. These authors contributed equally (Nathan R. Tucker, Mark Chaffin). Members of the Human Cell Atlas Lung Biological Network at provided in the Supplementary Materials.

## Abstract

Coronavirus disease 2019 (COVID-19) is a global pandemic caused by a novel severe acute respiratory syndrome coronavirus-2 (SARS-CoV-2). SARS-CoV-2 infection of host cells occurs predominantly via binding of the viral surface spike protein to the human angiotensin-converting enzyme 2 (ACE2) receptor.^1^ Hypertension and pre-existing cardiovascular disease are risk factors for morbidity from COVID-19,^2^ and it remains uncertain whether the use of angiotensin converting enzyme inhibitors (ACEi) or angiotensin receptor blockers (ARB) impacts infection and disease. Here, we aim to shed light on this question by assessing *ACE2* expression in normal and diseased human myocardial samples profiled by bulk and single nucleus RNA-seq.

---

The predominant portal of entry for SARS-CoV-2 is via *ACE2* expression in the upper respiratory epithelium and lungs,^3^ yet ACE2 is also abundantly expressed in the intestine, kidney, heart and testes. The observation that twenty percent of hospitalized patients with COVID-19 in Wuhan, China had evidence cardiac injury resuggest a possible pathological role for myocardial *ACE2* expression.^4^ In a single-center report of 416 patients hospitalized with COVID-19, 19.7% had evidence of cardiac injury. Moreover, the presence of cardiac injury was associated with a 5-fold increase in mechanical ventilation and a 51.2% mortality rate. In this context, the potential for a primary viral myocarditis with SARS-CoV-2 is gaining support. Prior findings of viral RNA from SARS-CoV-1, which also infects cells via ACE2, in 35% of autopsied hearts during an earlier SARS-CoV epidemic, lends credence to this possibility.^5^

Involvement of *ACE2* in COVID-19 pathogenesis has led to consideration of whether inhibitors of angiotensin converting enzyme (ACEi) and angiotensin receptor blockers (ARB) might modulate disease progression through increased expression of *ACE2*.^6^ Indeed, trials proposing to test the impact of both initiation (NCT04312009) and withdrawal of these agents are being advanced during the current pandemic. This uncertainty has provoked public statements by three leading cardiovascular societies advising continuation of these agents in the absence of compelling new data.^7^

To address these questions, we assessed *ACE2* expression in normal and diseased human myocardial samples from the Penn Human Heart Tissue Biobank by bulk and single nucleus RNA-seq. Our group has previously utilized this approach to study the transcriptional diversity of the cell subtypes and associated gene programs in the normal human heart.^8^

*ACE2* expression was assessed in the left ventricles of 11 individuals with dilated cardiomyopathy (DCM), 15 individuals with hypertrophic cardiomyopathy (HCM), and 16 non-failing controls defined as individuals without any overt indication of cardiac dysfunction. The baseline characteristics for the study population are provided in **Supplementary Table 1**. In an exploratory analysis, the effects of ACEi treatment on *ACE2* expression were evaluated in the subset of patients with HCM, in which six individuals were receiving ACEi and eight that were not prescribed an ACEi.

*ACE2* expression in the heart is strongest in pericytes, which line the microvasculature, but is also appreciable in vascular smooth muscle cells, fibroblasts and cardiomyocytes (**Fig. 1a**). We also evaluated the expression of the proteases encoded by *TMPRSS2* and *CTSL*, which facilitate membrane fusion and viral uptake during SARS-CoV-*2* infection.^1,9^ *TMPRSS2* was minimally expressed across all cell types, while *CTSL* displayed low levels of expression in all cell types with strongest expression in fibroblasts (18.1% and 56.7% of cells in the two populations found in non-failing controls), macrophages (39.1%), adipocytes (41.5%), and cardiomyocytes (11.4%) (**Supplementary Fig. 1**). These results are in concordance with recently released single nuclei data from Chen et al.^10^

**Figure 1:**
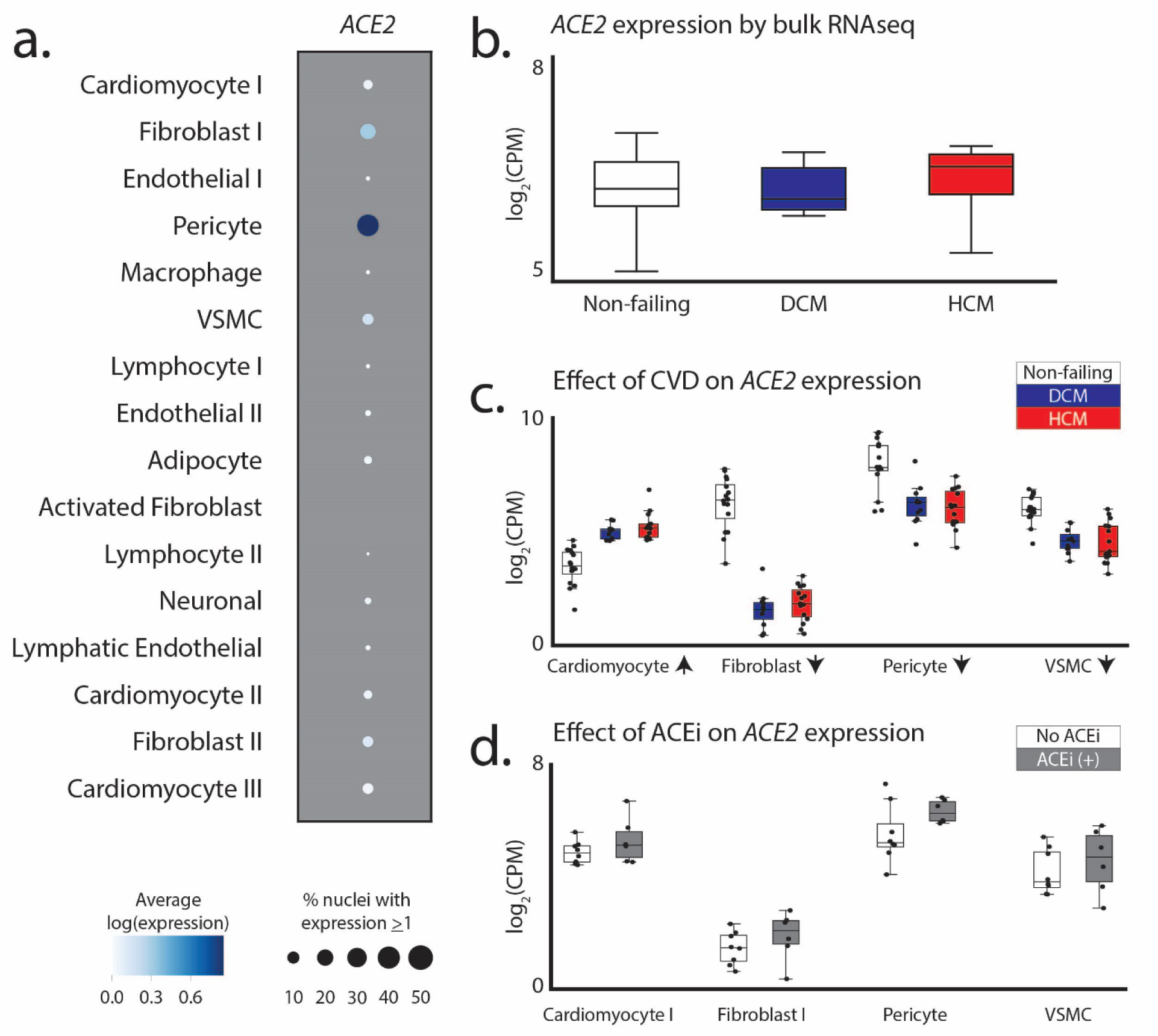
Assessment of *ACE2* expression in the human myocardium. **a**. Dot plot representing the relative expression of *ACE2* in non-failing left ventricles. Size and hue of the dot indicates the percent of nuclei expressing and mean log transformed counts for all nuclei in each cell type. VSMC: vascular smooth muscle cells **b**. Expression of *ACE2* by bulk RNA-Seq from non-failing, DCM and HCM ventricles. **c**. Single nucleus RNA-Seq from same tissue samples as (B) with mean expression of *ACE2* in cell subtypes with appreciable expression. Arrows indicate the direction of statistically significant changes when comparing non-failing to both DCM and HCM. **d**. Effects of ACEi on *ACE2* expression across cell types in HCM individuals. Boxes represent the 25-75% and whiskers represent the min-max range excluding outliers.

Analyses of previously generated bulk RNA-seq data from these individuals show no significant alterations in *ACE2* expression in the context of dilated or hypertrophic cardiomyopathy compared to non-failing controls (**Fig. 1b**). However, single nucleus RNA-seq highlighted a stark downregulation of *ACE2* expression in fibroblasts, pericytes, and vascular smooth muscle. There was a concomitant upregulation in ACE2 expression in cardiomyocytes in DCM and HCM (**Fig.1c**). The complete cell subtype differential expression analysis of *ACE2* in DCM and HCM compared to non-failing controls is provided in **Supplementary Tables 2 and 3**, respectively. A sensitivity analysis comparing the expression of *ACE2* in left ventricles samples from individuals with hypertrophic cardiomyopathy compared to non-failing controls after exclusion of individuals on ACE inhibitors or ARBs is provided in **Supplementary Table 4**.

Since patients with DCM were on an ACEi and nonfailing controls were not on an ACEi, our ability to examine effects of ACEi use on *ACE2* expression was limited to the the patients with HCM. Among these patients, there was a trend towards increased *ACE2* expression with ACEi treatment in all cell types (**Fig. 1d** and **Supplementary Table 5**), but these results were not statistically significant. Future studies with larger patient cohorts to examine cell type specific *ACE2* expression in additional disease states and in nonfailing donors receiving an ACEi or ARB are urgently needed. Further, it is important to note that transcriptional assessments do not reflect the effects of RNA half-life, translation efficiency, or protein turnover. As such, it will be important that surface localized ACE2 protein is also evaluated in future studies.

In conclusion, these data suggest that prior cardiovascular disease is a predominant driver of cardiomyocyte-specific increased transcription of *ACE2*. These findings may provide a pathological link for SARS-CoV-associated viral myocarditis.

## Data Availability

Given the urgent nature of the current pandemic, this analysis was confined to the genes known to interact with COVID-19 as described above. The cell subtype expression levels for each of the genes described in this analysis will be available on the Single Cell Portal at the Broad Institute (https://singlecell.broadinstitute.org/single_cell) upon publication. The description of the expression changes observed between dilated cardiomyopathy, hypertrophic cardiomyopathy and non-failing controls will be the basis of a distinct analysis, and the full snRNAseq dataset from these samples will be released upon publication of that manuscript.

## References

1. Hoffmann, M. et al. SARS-CoV-2 Cell Entry Depends on ACE2 and TMPRSS2 and Is Blocked by a Clinically Proven Protease Inhibitor. Cell (2020). doi:10.1016/j.cell.2020.02.052

2. Guan, W. et al. Clinical Characteristics of Coronavirus Disease 2019 in China. N. Engl. J. Med. NEJMoa2002032 (2020). doi:10.1056/NEJMoa2002032

3. Letko, M., Marzi, A. & Munster, V. Functional assessment of cell entry and receptor usage for SARS-CoV-2 and other lineage B betacoronaviruses. Nat. Microbiol. 5, 562–569 (2020).

4. Shi, S. et al. Association of Cardiac Injury With Mortality in Hospitalized Patients With COVID-19 in Wuhan, China. JAMA Cardiol. (2020). doi:10.1001/jamacardio.2020.0950

5. Booth, C. M. et al. Clinical Features and Short-term Outcomes of 144 Patients with SARS in the Greater Toronto Area. J. Am. Med. Assoc. 289, 2801–2809 (2003).

6. Clerkin, K. J. et al. Coronavirus Disease 2019 (COVID-19) and Cardiovascular Disease.Circulation (2020). doi:10.1161/CIRCULATIONAHA.120.046941

7. Bozkurt, B. (HFSA), Kovacs, R. (ACC) & Harrington, R. (AHA). HFSA/ACC/AHA statement addresses concerns re: using RAAS antagonists in COVID-19. Available at: https://professional.heart.org/professional/ScienceNews/UCM_505836_HFSAACCAHAstatement-addresses-concerns-re-using-RAAS-antagonists-in-COVID.jsp. (Accessed: 2nd April 2020)

8. Tucker, N. R. et al. Transcriptional and Cellular Diversity of the Human Heart. bioRxiv2020.01.06.896076 (2020). doi:10.1101/2020.01.06.896076

9. Simmons, G. et al. Inhibitors of cathepsin L prevent severe acute respiratory syndrome coronavirus entry. Proc. Natl. Acad. Sci. U. S. A. 102, 11876–11881 (2005).

10. Chen, L., Li, X., Chen, M., Feng, Y. & Xiong, C. The ACE2 expression in human heart indicates new potential mechanism of heart injury among patients infected with SARS-CoV-2. Cardiovasc. Res. (2020). doi:10.1093/cvr/cvaa078

